# Sociodemographic Differences in Population-Level Immunosenescence in Older Age

**DOI:** 10.1101/2022.03.05.22271952

**Authors:** Grace A. Noppert, Rebecca C. Stebbins, Jennifer Beam Dowd, Allison E. Aiello

## Abstract

**Background:** The COVID-19 pandemic has highlighted the urgent need to understand variation in immunosenescence at the population-level. Thus far, population patterns of immunosenescence are not well described.

**Methods:** We characterized measures of immunosenescence from newly released venous blood data from the nationally representative U.S Health and Retirement Study (HRS) of individuals ages 56 years and older.

**Findings:** Median values of the CD8+:CD4+, EMRA:Naïve CD4+ and EMRA:Naïve CD8+ ratios were higher among older participants and were lower in those with additional educational attainment. Generally, minoritized race and ethnic groups had immune markers suggestive of a more aged immune profile: Hispanics had a CD8+:CD4+ median value of 0.37 (95% CI: 0.35, 0.39) compared to 0.30 in Whites (95% CI: 0.29, 0.31). Blacks had the highest median value of the EMRA:Naïve CD4+ ratio (0.08; 95% CI: 0.07, 0.09) compared to Whites (0.03; 95% CI: 0.028, 0.033). In regression analyses, race/ethnicity and education were associated with large differences in the immune ratio measures after adjustment for age and sex. For example, each additional level of education was associated with roughly an additional decade of immunological age, and the racial/ethnic differences were associated with two to four decades of additional immunological age.

**Interpretation:** Our study provides novel insights into population variation in immunosenescence. This has implications for both risk of age-related disease and vulnerability to novel pathogens (e.g., SARS-CoV-2).

**Funding:** This study was partially funded by the U.S. National Institutes of Health, National Institute on Aging R00AG062749. AEA and GAN acknowledge support from the National Institutes of Health, National Institute on Aging R01AG075719. JBD acknowledges support from the Leverhulme Trust (Centre Grant) and the European Research Council grant ERC-2021-CoG-101002587

**Research in context:** *Evidence before this study:* Alterations in immunity with chronological aging have been consistently demonstrated across human populations. Some of the hallmark changes in adaptive immunity associated with aging, termed immunosenescence, include a decrease in naïve T-cells, an increase in terminal effector memory cells, and an inverted CD8:CD4 T cell ratio. Several studies have shown that social and psychosocial exposures can alter aspects of immunity and lead to increased susceptibility to infectious diseases.

*Add value of this study:* While chronological age is known to impact immunosenescence, there are no studies examining whether social and demographic factors independently impact immunosenescence. This is important because immunosenescence has been associated with greater susceptibility to disease and lower immune response to vaccination. Identifying social and demographic variability in immunosenescence could help inform risk and surveillance efforts for preventing disease in older age. To our knowledge, we present one of the first large-scale population-based investigations of the social and demographic patterns of immunosenescence among individuals ages 50 and older living in the US. We found differences in the measures of immunosenescence by age, sex, race/ethnicity, and education, though the magnitude of these differences varied across immune measures and sociodemographic subgroup. Those occupying more disadvantaged societal positions (i.e., minoritized race and ethnic groups and individuals with lower educational attainment) experience greater levels of immunosenescence compared to those in less disadvantaged positions. Of note, the magnitude of effect of sociodemographic factors was larger than chronological age for many of the associations.

*Implications for practice or policy and future research:* The COVID-19 pandemic has highlighted the need to better understand variation in adaptive and innate immunity at the population-level. While chronological age has traditionally been thought of as the primary driver of immunological aging, the magnitude of differences we observed by sociodemographic factors suggests an important role for the social environment in the aging human immune system.

## Introduction

Ageing of the immune system (i.e., immunosenescence) is hypothesized to contribute to the etiology of many age-related diseases including cardiovascular diseases, cancers, type 2 diabetes mellitus, neurodegenerative diseases, frailty, and premature mortality.^1-4^ Immunosenescence has also been implicated in reduced immune response to vaccination among ageing populations.^5^ While there is growing evidence that differences in immune status may underlie many age-related diseases, population patterns of immunosenescence in older age have not been well described.

As individuals age, several functional changes occur in both the innate and adaptive immune systems including dysfunction in the T cell compartment,^6-9^ increasing chronic low-grade inflammation (i.e., ‘inflammaging’) and diminished ability to fight infections.^10^ Alterations to the innate immune system observed in older age include decreased leukocyte production and function, reduced phagocytic capability of neutrophils, elevated monocytes and macrophages, and a functional shift towards a proinflammatory phenotype. The thymus gland, which plays a key role in the adaptive immune response, undergoes chronic involution after puberty, resulting in a continuous decline in T cell output with age and differing rates of involution by sex.^11^ In addition, T cell ratios and cell surface markers change over time, including an inversion of CD4+:CD8+ T cell ratios, a decrease in naïve T cells, and an accumulation of effector memory T cells with limited function.^12^ Together, these age-related immune alteration form a phenotype of immunosenescence.

Concurrently, antigenic stimulation across the life course drives expansion of memory and effector T cell subsets contributing to an increase in the number of pathogen-specific, highly differentiated T cells.^13^ It is thought that cytomegalovirus (CMV) infection, in particular, may be a key driver of these trends.^14,15^ CMV is a highly prevalent herpesvirus infection in older age populations and has been implicated as a major driver of age-associated alterations to the T cell repertoire^16^ with expansion of T cells specific for CMV and decreases in T cells specific to other pathogens over time.^17^

Despite evidence that changes in immunity contributes to many of the hallmark diseases of ageing, prior research has relied on small, often non-representative samples (e.g., clinical cohorts) and investigated individual markers rather than employing multiple indicators across the immune system. Using newly released venous blood data collected in 2016 from the population-based U.S. Health and Retirement Study (HRS), this study characterizes markers and ratios of T-cell immunosenescence in a population of individuals ages 56 years and older. We also examined other age-related immune cell markers of the adaptive (B-cells, Natural Killer cells) and innate (monocytes, lymphocytes, Natural Killer cells) immune system. We assessed differences in these markers of immunosenescence by sociodemographic characteristics including age, sex, race/ethnicity, and educational attainment. As one of the first population-based studies of immunosenescence, our study provides novel insights into population variation in immunity, with implications for age-related disease, vaccine response, and vulnerability to novel pathogens, such as SARS-CoV-2.

## Methods

### Study Population

Data come from the U.S. Health and Retirement Study (HRS), the largest on-going nationally representative longitudinal survey of older adults in the U.S. HRS began in 1992 and included over 22,000 adults over the age of 50 years at baseline and interviewed every two years. HRS survey design and methods have been described previously.^18^ Data collection consisted of face-to-face baseline interviews and primarily telephone interviews for follow-up waves, until 2006, when half the sample (alternated at each subsequent wave) was randomly assigned face-to-face interviews to enhance physical and biological measures. Our analysis uses existing sociodemographic data from the 2016 core interview, with immune data from the 2016 Venous Blood Study.^19^ Of the 9,934 participants who consented to the venous blood draw and were tested (consent rate=78.5%, completion rate=65%), our analysis included individuals who 1) had positive survey weights, 2) full covariate data, and 3) had one valid immune biomarker measure such that we were able to calculate at least one ratio measure. We also excluded two individuals with extreme values on two of the ratio measures.

### Immune Measures

Immunophenotyping was performed on the entire VBS sample. Further details on blood sample processing, laboratory assays, and the characterization of subtypes of T, B, monocytes, and natural killer (NK) cells are described in Thyagarajan et al., 2021.^8^ We focused on 10 specific cell subtypes: CD4+ (CD3+ CD19- CD8- CD4+), naïve CD4+ (CD45RA+ CD28- CD95-), terminally differentiated effector memory (EMRA) CD4+ (CD3+ CD19- CD8- CD4+ CD45RA+ CCR7- CD28-), CD8+ (CD3+ CD19- CD8+ CD4), Naïve CD8+ (CD3+ CD19- CD8+ CD4- CD45RA+ CCR7+ CD28+), EMRA CD8+ (CD3+ CD19- CD8+ CD4- CD45RA+ CCR7- CD28-), IgD+ Memory B Cells (CD3- CD19+ IgD+ CD27+), IgD- naïve B Cells (CD3-CD19+ IgD- CD27+), NK Cells: CD56 Low (CD3- CD19- CD20- CD14- CD16+ CD56LO), and NK Cells: CD56 High (CD3- CD19- CD20- CD14- CD16+ CD56HI).

For the primary analyses, we used a continuous percentage measure for each cell type. The percentages were defined in terms of the proportion of the parent cell population. For example, the percentage of naïve CD4+ T cells is defined as the proportion of naïve CD4+ T cells out of the total CD4+ cell parent population.

We utilized several T-cell markers to create ratios measures of immunosenescence. CD4+:CD8+ T cell ratios have been widely used in clinical research to indicate immunocompromised health and this ratio known to decrease with age.^20^ In addition, we followed a previously published methods for creating ratios of T cell immunosenescence, developed by Aiello et al.^21,22^ Following these approaches, we assessed the following ratios:

A. CD8+:CD4+
B. EMRA CD4+:Naïve CD4+
C. EMRA CD8+:Naïve CD8+

We also developed three novel ratio measures that have been less utilized in studies of immunosenescence and provide insights on both adaptive and innate immune changes with age:

D. Memory:Naïve B Cell (adaptive)
E. NK Cells CD56 Low to CD56 High (adaptive and innate)
F. Monocyte: Lymphocyte (innate)

The analyses using these novel ratio measures are included in the supplementary material. For full details on which variables were used to create each ratio measure, see **Supplemental Material**.

We added a small constant 0.001 to all cell types to avoid dividing by a zero value, and all ratio measures were natural-log transformed prior to inclusion in regression models in order to account for skewness.

### Cytomegalovirus

In addition, we examined whether continuous CMV IgG antibody levels mediate the relationship between sociodemographic factors and immunosenescence. The role of CMV in immunosenescence remains unclear. This large DNA virus may drive immunosenescence or interact biologically with the immune system in ways that accelerate immunosenescence. Alternatively, immunosenescence may lead to loss of immune control over CMV and subclinical reactivation. Importantly, CMV has known associations with socioeconomic factors at the population-level.^21,23-26^ CMV immunoglobulin (Ig) G levels were measured in serum using the Roche e411 immunoassay analyzer.

### Sociodemographic Variables

Self-reported sociodemographic characteristics including age, sex, race/ethnicity, and educational attainment were collected in the 2016 core interview. Educational attainment was categorized as less than a high school diploma, high school diploma, some college, or a college diploma and higher. Race/ethnicity was categorized as non-Hispanic White, non-Hispanic Black, Hispanic, or Other Race, and sex was self-reported as either male or female.

### Statistical Analyses

We calculated descriptive statistics for the study population overall and stratified by age group, sex, race/ethnicity, and educational attainment. We compared the median values and 95% confidence interval of each of the immune ratios by sociodemographic characteristic.

We ran a series of linear regression models to estimate the association between each sociodemographic characteristic and the three primary immune measures. **Model 1** included age and sex. **Model 2** included age, sex, and educational attainment. **Model 3** included age, sex, and race/ethnicity. **Model 4** included age, sex, race/ethnicity, and educational attainment. Additionally, to investigate whether CMV partially mediates the association between the sociodemographic characteristics and immune outcomes, **Model 5** includes all covariates plus CMV IgG levels. The regression models estimate the change in the mean value of the log-transformed immune ratio measure associated with each category of sociodemographic group compared to a referent.

All analyses were weighted with the 2016 VBS weights and the statistical models accounted for the complex survey design. Statistical analyses were conducted in SAS 9.4 (SAS Institute, Inc., Cary, North Carolina), and figures were generated in R.

## Results

### Sample Characteristics

Overall, the sample had a median age of 66 years and was 54% female. The majority of the sample was Non-Hispanic White (78%) and had achieved greater than a high school education. The median CD8+:CD4+ ratio was 0.318 (95% CI: 0.311, 0.325), the median of the EMRA:Naïve CD4+ ratio was 0.037 (95% CI: 0.035, 0.039), and the median EMRA: Naive CD8+ ratio was 2.23 (95% CI: 2.13, 2.34). Finally, 66% of the study population was CMV seropositive, and among those, the median CMV IgG antibody levels were 384.27 (95% CI: 367.12, 401.42). Full demographic characteristics are given in **Table 1**.

**Table 1.**
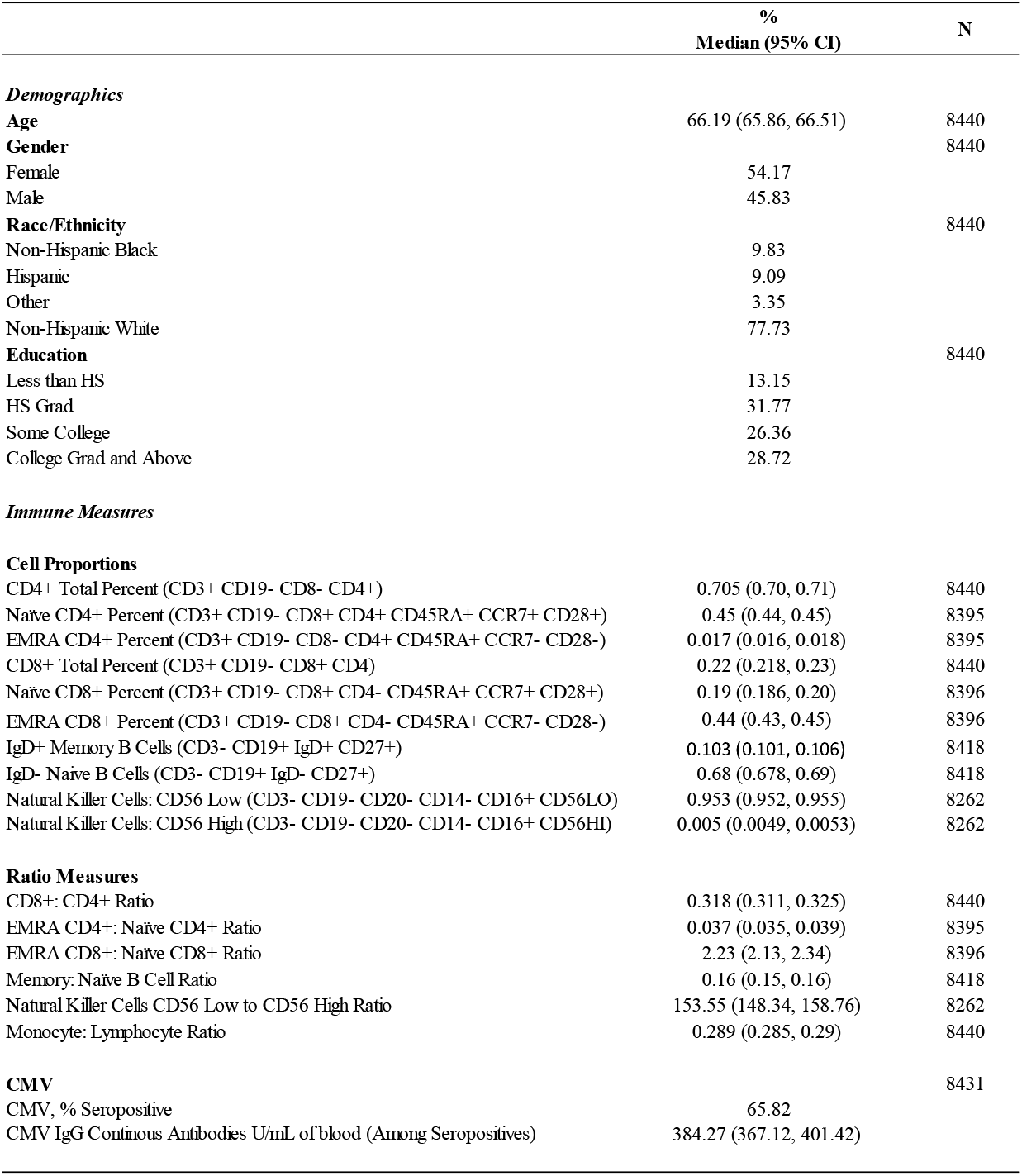
Demographic characteristics of the Health & Retirement Study analytic sample.

### Distribution of Immune Measures by Sociodemographic Characteristics

We compared the median values of each immune measure across age group, sex, race/ethnicity, and education (**Table 2 and Figure 1**). As expected, the median values of the CD8+:CD4+, EMRA:Naive CD4+, and EMRA:Naive CD8+ ratios increased with age. For example, among those 56-65 years, the median CD8+:CD4+ value was 0.31 (95% CI: 0.29, 0.32) compared to the median of 0.37 (95% CI: 0.33, 0.41) among those 86+ years. Comparing the median values of the EMRA:Naive CD8+ ratio, those 56-65 years old had a median value of 1.44 (95% CI: 1.30, 1.57) compared to those 86+ with a median value of 6.71 (95% CI: 5.53, 7.88).

**Table 2.** Distribution of immune measures by age group, sex, race/ethnicity and educational attainment.

|  | Age Group |  |  |  | Sex |  |
| --- | --- | --- | --- | --- | --- | --- |
|  | 56-65 | 66-75 | 76-85 | 86+ | Female | Male |
|  | Median (95% CI) | Median (95% CI) | Median (95% CI) | Median (95% CI) | Median (95% CI) | Median (95% CI) |
| CD8+: CD4+ Ratio | 0.31 (0.29, 0.32) | 0.31 (0.30, 0.33) | 0.35 (0.34, 0.36) | 0.37 (0.33, 0.41) | 0.30 (0.29, 0.31) | 0.34 (0.33, 0.35) |
| EMRA CD4+: Naïve CD4+ Ratio | 0.03 (0.027,.033) | 0.037 (0.035,.04) | 0.05 (0.04,.05) | 0.06 (0.05,.07) | 0.038 (0.035, 0.040) | 0.035 (0.032,.038) |
| EMRA CD8+: Naïve CD8+ Ratio | 1.44 (1.30, 1.57) | 2.49 (2.29,2.70) | 4.39 (4.09,4.69) | 6.71 (5.53, 7.88) | 1.96 (1.79, 2.12) | 2.56 (2.39, 2.74) |
| Memory: Naïve B Cell Ratio | 0.16 (0.15,.17) | 0.15 (0.14,.16) | 0.16 (0.15,.18) | 0.19 (0.16,.22) | 0.15 (0.147,0.16) | 0.17 (0.16,.18) |
| Natural Killer Cells CD56 Low to CD56 High Ratio | 142.24 (134.55,149.93) | 154.96 (147.00,162.71) | 176.98 (168.51, 185.45) | 176.92 (153.58, 200.26) | 146.32 (139.86, 152.79) | 160.34 (153.51, 167.16) |
| MLR | 0.27 (0.26,.273) | 0.29 (0.284,.30) | 0.33 (0.324,.34) | 0.35 (0.33,.37) | 0.27 (0.26,.271) | 0.32 (0.31,.32) |
| CMV, % Seropositive | 59.46 | 67.08 | 75.21 | 80.92 | 70.83 | 59.9 |
| CMV IgG Continous Antibodies U/mL of blood (Among Seropositives) | 400.83 (369.47, 432.20) | 373.62 (340.18, 407.07) | 380.38 (356.24, 404.53) | 364.09 (290.96, 437.22) | 431.54 (406.01, 457.28) | 327.17 (303.45, 350.90) |

|  | Race/Ethnicity |  |  |  |
| --- | --- | --- | --- | --- |
|  | Non-Hispanic Black | Hispanic | Other Race | Non-Hispanic White |
|  | Median (95% CI) | Median (95% CI) | Median (95% CI) | Median (95% CI) |
| CD8+: CD4+ Ratio | 0.35 (0.33, 0.37) | 0.37 (0.35, 0.39) | 0.35 (0.31, 0.39) | 0.30 (0.29, 0.31) |
| EMRA CD4+: Naïve CD4+ Ratio | 0.08 (0.07, 0.09) | 0.07 (0.06,.08) | 0.04 (0.03,.05) | 0.03 (0.028,.033) |
| EMRA CD8+: Naïve CD8+ Ratio | 1.84 (1.66, 2.02) | 3.42 (3.12,3.71) | 2.18 (1.77, 2.58) | 2.16 (2.03, 2.28) |
| Memory: Naïve B Cell Ratio | 0.19 (0.18, 0.21) | 0.16 (0.14,.17) | 0.12 (0.1,.15) | 0.16 (0.15,.16) |
| Natural Killer Cells CD56 Low to CD56 High Ratio | 203.10 (188.89, 217.30) | 160.51 (147.01, 174.01) | 179.05 (161.54, 196.56) | 144.35 (139.38, 149.32) |
| MLR | 0.25 (0.24, 0.25) | 0.25 (0.24,.26) | 0.25 (0.23,0.26) | 0.300 (0.296, 0.304) |
| CMV, % Seropositive | 88.33 | 93.42 | 84.85 | 58.93 |
| CMV IgG Continous Antibodies U/mL of blood (Among Seropositives) | 435.72 (387.51, 483.94) | 401.29 (361.52, 441.05) | 327.82 (245.70, 409.95) | 373.86 (352.74, 394.97) |

|  | Education |  |  |  |
| --- | --- | --- | --- | --- |
|  | Less than HS<br>Median (95% CI) | HS Grad<br>Median (95% CI) | Some College<br>Median (95% CI) | College +<br>Median (95% CI) |
| CD8+: CD4+ Ratio | 0.39 (0.36, 0.41) | 0.32 (0.31, 0.33) | 0.32 (0.30, 0.33) | 0.28 (0.27, 0.29) |
| EMRA CD4+: Naïve CD4+ Ratio | 0.064 (0.06, 0.07) | 0.04 (0.037,0.044) | 0.03 (0.029, 0.034) | 0.027 (0.02,0.03) |
| EMRA CD8+: Naïve CD8+ Ratio | 3.53 (3.21, 3.840) | 2.4 (2.18,2.63) | 1.96 (1.79, 2.13) | 1.86 (1.67,2.05) |
| Memory: Naïve B Cell Ratio | 0.17 (0.16, 0.19) | 0.15 (0.14,.16) | 0.17 (0.16,0.18) | 0.16 (0.15,0.17) |
| Natural Killer Cells CD56 Low to CD56 High Ratio | 177.83 (167.44, 188.23) | 160.28 (150.38,170.18) | 147.42(137.22,157.63) | 141.94 (134.36,149.51) |
| MLR | 0.26 (0.25, 0.27) | 0.29 (0.285,0.30) | 0.29 (0.28,0.293) | 0.297 (0.289, 0.31) |
| CMV, % Seropositive | 87.98 | 69.68 | 62.65 | 54.35 |
| CMV IgG Continuous Antibodies U/mL of blood (Among Seropositives) | 437.13 (400.38, 474.89) | 396.02 (367.22, 424.82) | 373.38 (342.58, 404.19) | 340.27 (315.63, 264.91) |

**Figure 1.**
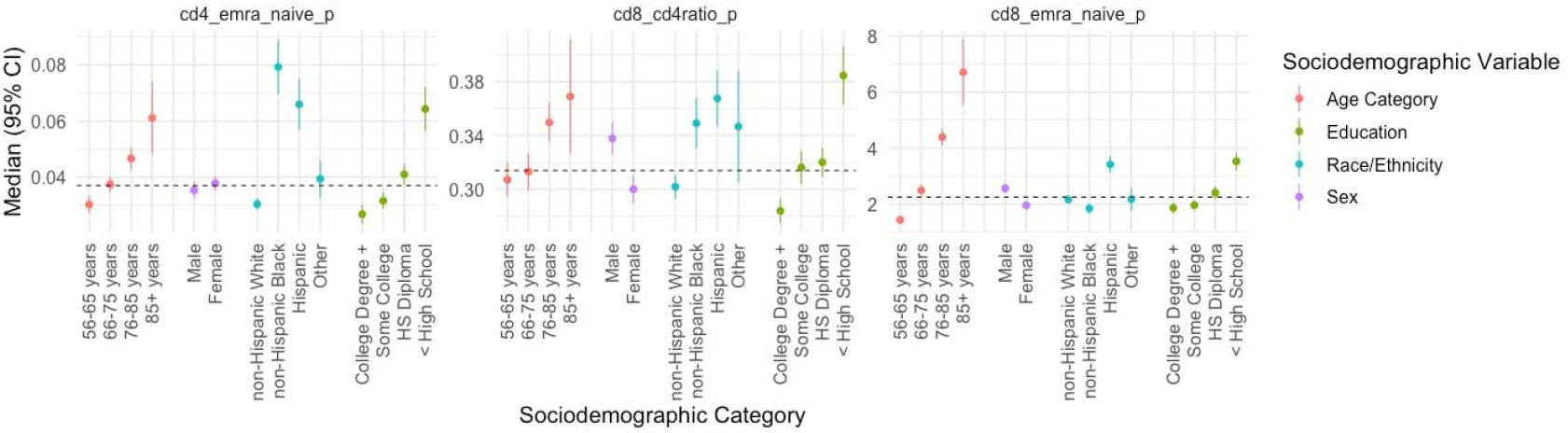
Distribution of each immune measure by age category, sex, race/ethnicity, and educational attainment. Medians and 95% confidence intervals are displayed. **Panel A** corresponds to the CD4+ EMRA:Naïve ratio. **Panel B** corresponds to the CD8+:CD4+ ratio. **Panel C** corresponds to the CD8+ EMRA:Naïve ratio.

We also observed sex differences in the median value of each of the outcome measures. Females had lower median values for the CD8+:CD4+ and EMRA:Naive CD8+ ratios compared to men. For the CD8+:CD4+ ratio men had a median value of 0.34 (95% CI: 0.33, 0.35) compared to the female median of 0.30 (95% CI: 0.29, 0.31), and for the EMRA:Naive CD8+ men had a median value of (2.56; 95% CI: 2.39, 2.74) compared to females with a median of 1.96 (95% CI: 1.79, 2.12).

The median values of each of the immune outcomes also differed across race/ethnic group though the pattern depended on the particular immune markers. Overall, minoritized racial ethnic subgroups had immune markers suggestive of a more aged profile compared to Non-Hispanic Whites. For example, Hispanics had the highest value of the CD8+:CD4+ ratio with a value of 0.37 (95% CI: 0.35, 0.39) compared to Non-Hispanic Whites with the lowest value of 0.30 (95% CI: 0.29, 0.31). For the EMRA:Naïve CD4+ ratio, Non-Hispanic Blacks had the highest median value of 0.08 (95% CI: 0.07, 0.09) compared to Non-Hispanic Whites with the lowest value of 0.03 (95% CI: 0.028, 0.033). However, for the EMRA:Naïve CD8+ ratio, Hispanics had the highest value of 3.42 (95% CI: 3.12, 3.71) while Non-Hispanic Blacks had the lowest value of 1.84 (95% CI: 1.66, 2.02).

Finally, we observed an educational gradient across all three immune measures with more education associated with lower values, suggesting a less aged immune profile. For example, those with less than a high school education had a median CD8+:CD4+ ratio of 0.39 (95% CI: 0.36, 0.41) compared to those with a college degree or higher with a value of 0.28 (95% CI: 0.27, 0.29). The same trend was observed for both the EMRA:Naïve CD4+ ratio and the EMRA:Naïve CD8+ ratio.

### Associations between sociodemographic factors and immune function

Figure 2. and **Tables S1-S3** show the associations between sociodemographic characteristics and the immune ratios measures, adjusted for covariates. In model 1, we found that for each additional year of age, there was a higher mean value of all three ratio measures CD8+:CD4+, CD4+ EMRA:Naïve ratio and CD8+ EMRA:Naïve ratio when adjusted for sex. For example, a one-year increase in age was associated with 0.01 (95% CI: 0.004, 0.01) higher CD8+:CD4+ ratio. There were also significant associations between sex and both the CD8+:CD4+ ratio and the CD8+ EMRA:Naïve ratio, with females having a lower mean value compared to males. For the CD8+:CD4+ ratio, females had a mean value 0.15 lower than males (95% CI: -0.19, -0.11) and for the CD8+ EMRA:Naïve females had a mean value 0.36 lower than males (95% CI: -0.43, -0.29). The CD4+ EMRA:Naïve ratio did not differ by sex.

**Figure 2.**
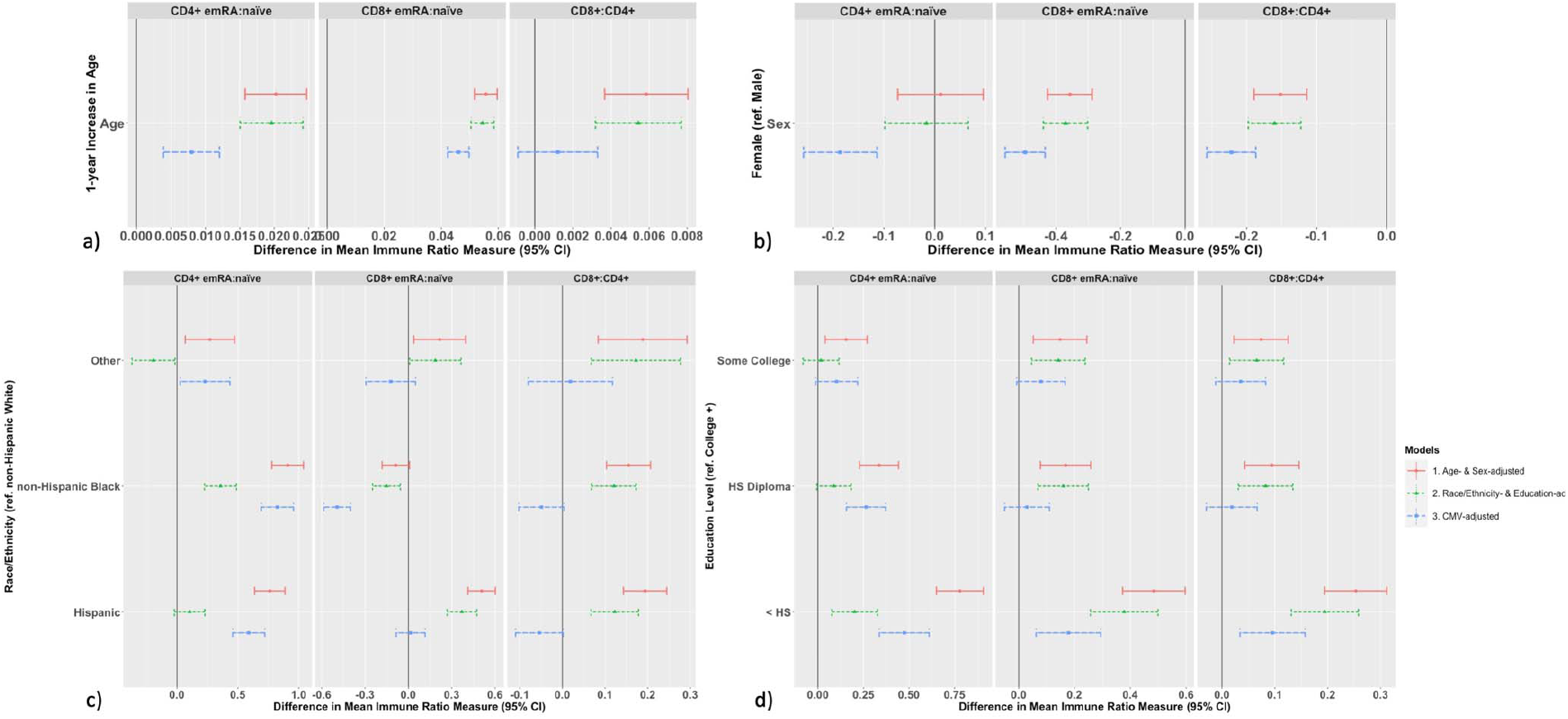
Results of the regression analyses estimating the associations between age (**Panel A**), sex (**Panel B**), race/ethnicity (**Panel C**), and educational attainment (**Panel D**) and each of the three immune outcomes. Beta coefficients and 95% confidence intervals are depicted.

Models 2 and 3 show the age- and sex-adjusted estimates for the associations between the immune ratio measures and education and race/ethnicity, respectively. For all three measures, higher educational attainment was associated with lower mean values on all immune measures, suggesting a less aged immune profile. The educational differences were most strongly observed for the EMRA:Naïve CD4+ ratio where those with less than a high school education had a mean value 0.78 (95% CI: 0.65, 0.91) higher than those with a college degree or higher while those with some college had a mean value 0.15 (95% CI: 0.04, 0.27) higher than those with a college degree.

With non-Hispanic White as the referent, all three immune ratios varied significant by racial and ethnic group. For all three ratio measures, a higher value is indicative of a more aged immune profile. For the CD8+:CD4+ ratio, this contrast was most strongly observed comparing Hispanics and those classified as “Other” race/ethnicity to Non-Hispanic Whites: both Hispanics and “Other” race/ethnicity had a mean value 0.19 (95% CI: 0.14, 0.24; 95% CI: 0.08, 0.29, respectively) higher compared to Non-Hispanic Whites. Non-Hispanic Blacks had a mean EMRA:Naïve CD4+ ratio 0.91 (95%CI:0.78,1.04) higher than non-Hispanic Whites. The difference between non-Hispanic Blacks and non-Hispanic Whites’ mean EMRA:Naïve CD8+ ratio was not significant, though Hispanics had an elevated value (0.51 (95% CI: 0.41, 0.60)). In general, these associations were slightly attenuated when race/ethnicity and education were included in the same regression model (Model 4). See **Figure 2, Tables S1-S3** for all relevant measures of association.

After adding CMV IgG levels to the model (Model 5) many of the education and race/ethnicity estimates, though not all, were attenuated, indicating that CMV may be a direct or indirect link between social factors and immune ageing.

To ease interpretation of the estimates, we calculated age-standardized estimates of each sociodemographic association by dividing the beta coefficients by the age coefficient in the race/ethnicity-, education-, age-, and sex-adjusted models. These estimates are represented as additional years of immune system ageing compared to the referent and displayed in **Figure 3**. We found that lower educational attainment or being a member of a minoritized racial or ethnic group was typically associated with additional years of immunological ageing. For example, compared to those with a college degree, those who completed less than a high school diploma had a CD4+ EMRA:Naïve ratio equivalent to 40 (95%CI: 33, 46) additional chronological years of immune ageing. Compared to non-Hispanic Whites, Hispanics had a CD8+:CD4+ ratio equivalent to roughly an additional 36 (95%CI: 26, 45) chronological years of immunological ageing, though the estimates for all ratio measures were attenuated when including education in the model.

**Figure 3.**
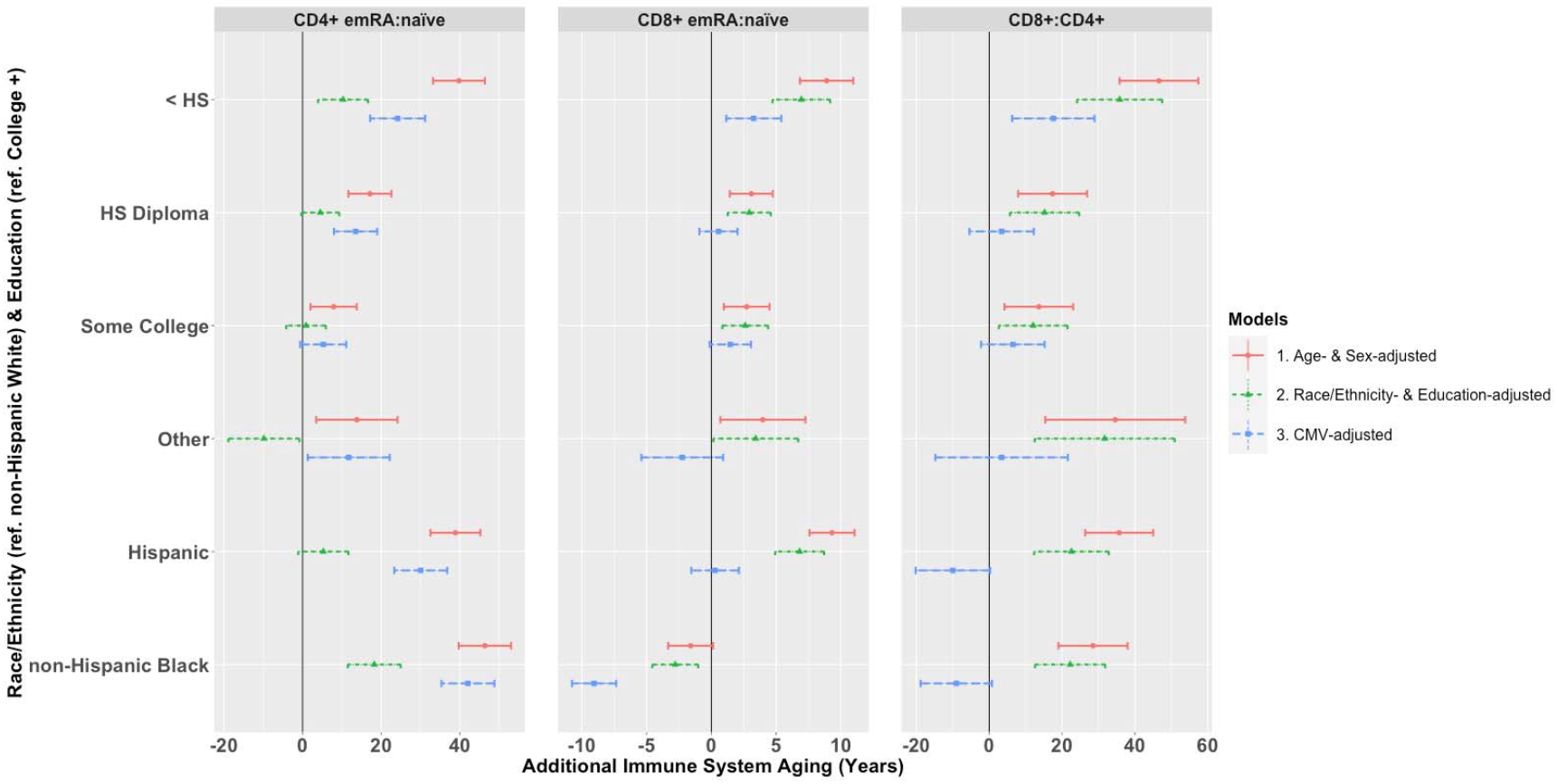
Results of the analyses estimating the additional years of immune system aging by strata of educational attainment (ref: College degree) and race/ethnicity (ref: non-Hispanic White). **Panel A** corresponds to the CD4+ EMRA:naïve ratio. **Panel B** corresponds to the CD8+ EMRA:naïve ratio. **Panel C** corresponds to the CD8+:CD4+ ratio.

We also replicated the analyses with the three additional immune ratio outcomes: memory:naïve B cell ratio, NK cells CD56 low: CD56 high ratio, and monocyte: lymphocyte ratio. Associations with sociodemographic characteristics were mixed and not in a consistent direction. With novel immune measures, we are cautious in interpreting how higher or lower values correspond to immunosenescence, but hope these estimates provide new descriptive information. Full results are available in supplemental **Figure S1-S3** and **Tables S4-S6**.

## Discussion

The large variation in patterns of COVID-19 disease susceptibility and vaccination response to SARS-CoV-2, has highlighted the critical need to better understand population differences in adaptive and innate immunity. We identified significant associations between sociodemographic factors and immunosenescence among individuals 56 years and older. We found differences in the levels of each immunosenescence ratio by age group, sex, race/ethnicity, and education, though the magnitude of the differences varied by immune measure and sociodemographic characteristic. Overall, those occupying more disadvantaged societal positions (i.e., minoritized race and ethnic groups and individuals with lower educational attainment) had indicators of more advanced immunosenescence compared to those in less disadvantaged positions. The demographic predictors of the immune measures we described here have been assessed in only a few other population-based studies and these were smaller sub-samples.^21,22^ Our results showed that sociodemographic factors, specifically race/ethnicity and education, were associated with large differences in immune measures even compared to those associated with chronological age. For example, each additional level of education was associated with roughly an additional decade of immunological age, and the racial/ethnic differences were about twice to four times that magnitude (i.e., two to four decades of additional immunological age; **Figure 3**). Finally, in our preliminary analyses, we found that inclusion of CMV in the models resulted in partial, and in some cases full, attenuation of the estimates between the sociodemographic characteristic and the immune outcome measures, similar to previous studies.^21,22^ Given the strong association of CMV with sociodemographic factors, more work on the causal role of CMV in these associations is warranted. Overall, these findings suggest an important role for the social environment in addition to chronological age in shaping immunosenescence at the population-level.

The social environment could accelerate immunological ageing via several pathways. Socially disadvantaged individuals are more likely to encounter both infectious and other immune-taxing exposures across their life course. For example, social differences in housing, transportation, and occupations may lead to more infectious exposures, highlighted by COVID-19.27 Living in disadvantaged neighborhoods can increase the quantity and frequency of exposure to pollution, noise, and psychosocial stressors that activate cortisol production via the HPA-axis, potentially interacting directly with immune cells.^28^ Indeed, previous research has shown that psychosocial trauma is associated with changes in these ratios in the same direction we identified, suggesting that psychosocial stressors may play a role in the association between disadvantage and immunosenescence.^22^ If stressors age the immune system, our findings could have key implications for understanding the biological mechanisms linking disadvantage to poor health. Further research explicating stress-related pathways to immunosenescence are warranted.

We observed higher levels of T cell immunosenescence in racial and ethnic minorities and those with less education. Importantly, the magnitudes of these sociodemographic differences are large in comparison to the differences observed by age. Since we would expect to detect differences only on the scale of a decade of chronological ageing (due to small changes and noise at the population-level), the roughly one to four decades of additional immunological ageing associated with social factors is striking. It is particularly important to consider the chronic, life course nature of these social exposures, which may help explain the magnitude of these associations. The weathering hypothesis, for example, posits that the chronic exposure to oppressive social environments explains part of the racial and ethnic disparities observed for health.^29^ To our knowledge, the potential role of immunosenescence in weathering and health disparities has not been thoroughly investigated.

While this study significantly advances our understanding of population-level immunosenescence, there are several key limitations that should be considered. First, the immune measures are only measured at a single point in time. These data were all collected as part of the 2016 VBS; no longitudinal data are available. Thus, we cannot make inferences regarding life course trajectories of immune biomarkers or differential speed of immunosenescence. Second, measurement of T cell phenotypes in large-scale cohort studies is logistically demanding, and there is likely to be technical error introduced through the time it takes to process samples and cell loss. These markers are not only sensitive to the conditions under which they are collected and processed, but also sensitive to human variation at the time of collection (e.g., may be altered in response to acute illness). While large and population-based, the HRS over-represents non-Hispanic Whites, thus our results are not fully generalizable to the U.S. population. There is also differential attrition by race/ethnicity, and our analytic sample only includes individuals who survived to age 56 and enrolled in the study. An additional limitation is the possibility of selection bias though it is difficult to disentangle the direction of the bias. Generally, those who agree to enroll in studies such as this are healthier than the average population and experience less life disadvantage.^30^ As a result, selection may be biasing associations between immunosenescence measures and sociodemographic variables. Future studies should investigate these relationships earlier in the life course, before individuals are dying of age-related diseases, to understand the extent of this bias. Despite using previously established methods for our T cell ratio measures, these ratios likely don’t capture the dynamic links between T cells, B cells, NK cells, macrophages, and lymphocytes. Development of measures that better capture interactions between these markers and how they evolve with ageing could improve our models of immunosenescence.

## Conclusion

Our study describes the relationship between sociodemographic factors and key measures of immunosenescence in a large, population-based cohort. We found evidence that lower educational attainment and minoritized racial ethnic status was associated with higher levels of immunosenescence. While age has traditionally been thought of as the primary driver of immune differences, the magnitude of differences by sociodemographic factors observed, suggests that the social environment also plays an important role in ageing the human immune system.

## Data Availability

The Health and Retirement Study date are publicly available through https://hrs.isr.umich.edu/. The Venous Blood Study data are restricted but can be accessed through a data use agreement with the Health and Retirement Study.

## Statements

### Author Contributions

Consent to submit has been received explicitly from all coauthors. Authors whose names appear on the submission have contributed sufficiently to the scientific work and therefore share collective responsibility and accountability for the results.

