## Supplement for "Sociodemographic Differences in Population-Level Immunosenescence in Older Age"

**Supplementary Material**

Section I: Development of Immunosenescence Ratio Measures ………………………………2

Section II: Supplemental Tables ………………………………………………………………. 3

Section III: Supplemental Figures ……………………………………………………………. 9

Section I: Development of Immunosenescence Ratio Measures

A. CD8+: CD4+ ratio = (PCD8T_PCT + **0.001)** / (PCD4T_PCT + **0.001)**
B. EMRA CD4+: Naïve CD4+ Ratio = (PCD4TEMRA_PCT + **0.001)** / (PCD4N_PCT + **0.001)**
C. EMRA CD8+: Naïve CD8+ Ratio = (PCD8TEMRA_PCT + **0.001)** / (PCD8N_PCT + **0.001)**
D. Memory: Naïve B Cell Ratio = (PIGD_PLUS_MEMB_PCT + **0.001**) / (PNAIVEB_PCT + **0.001**)
E. NK Cells CD56 Low to CD56 High Ratio = (PNKLO_PCT + **0.001**) / (PNKHI_PCT + **0.001**)
F. Monocyte: Lymphocyte Ratio = (PMONO + **0.001**) / (PLYMP + **0.001**)

Section II: Supplemental Tables

**Table S1. Regression Results for CD8+:CD4+ Ratio.**

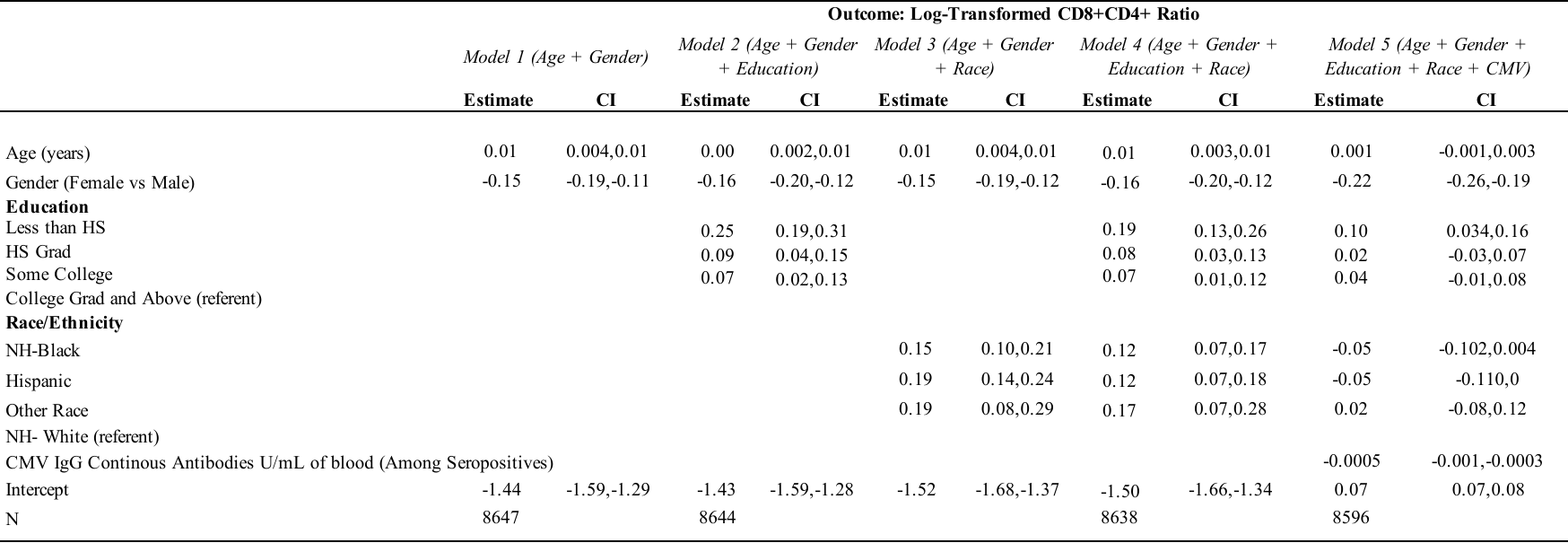

**Table S2. Regression Results for CD4+ EMRA:Naïve Ratio.**

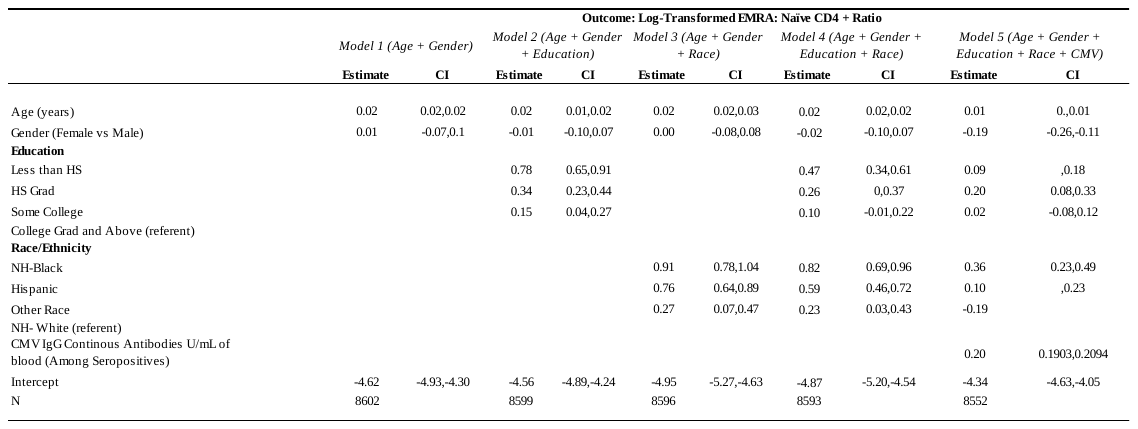

**Table S3. Regression Results for CD8+ EMRA:Naïve Ratio.**

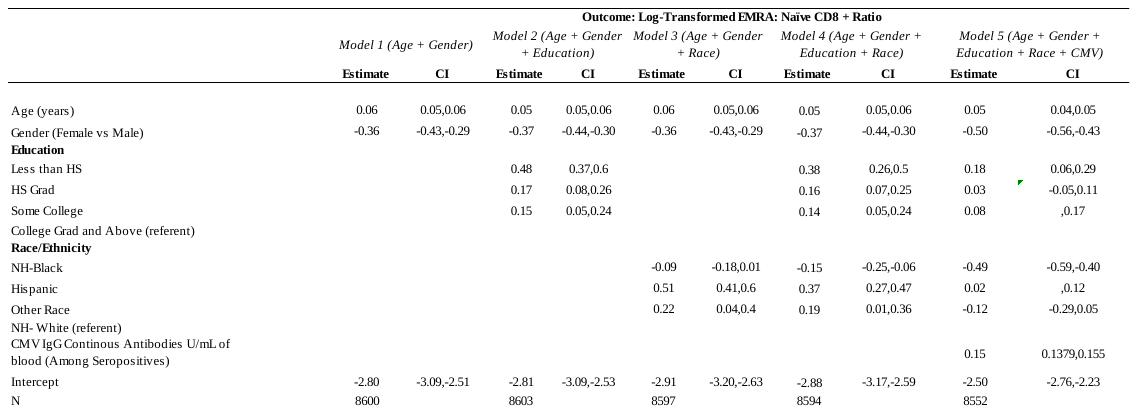

**Table S4. Regression Results for B Cells Ratio.**

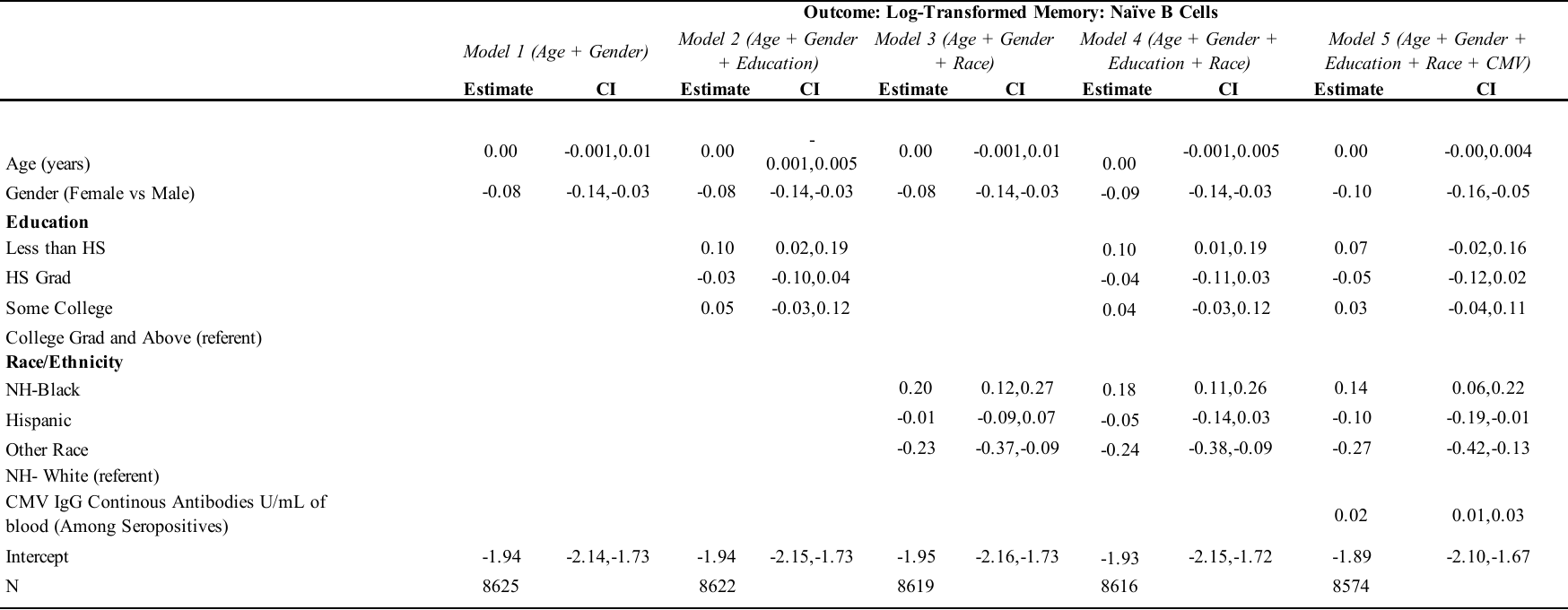

**Table S5. Regression Results for Natural Killer Cells Ratio.**

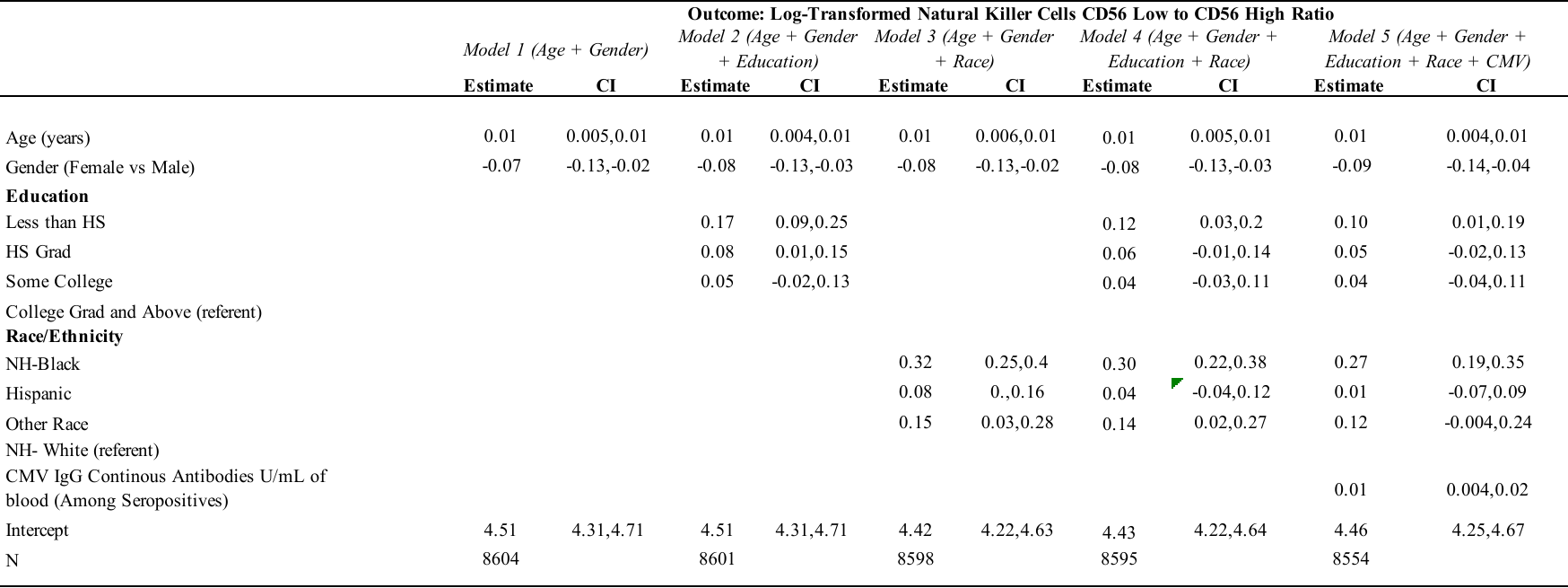

**Table S6. Regression Results for Monocyte:Lymphocyte Ratio.**

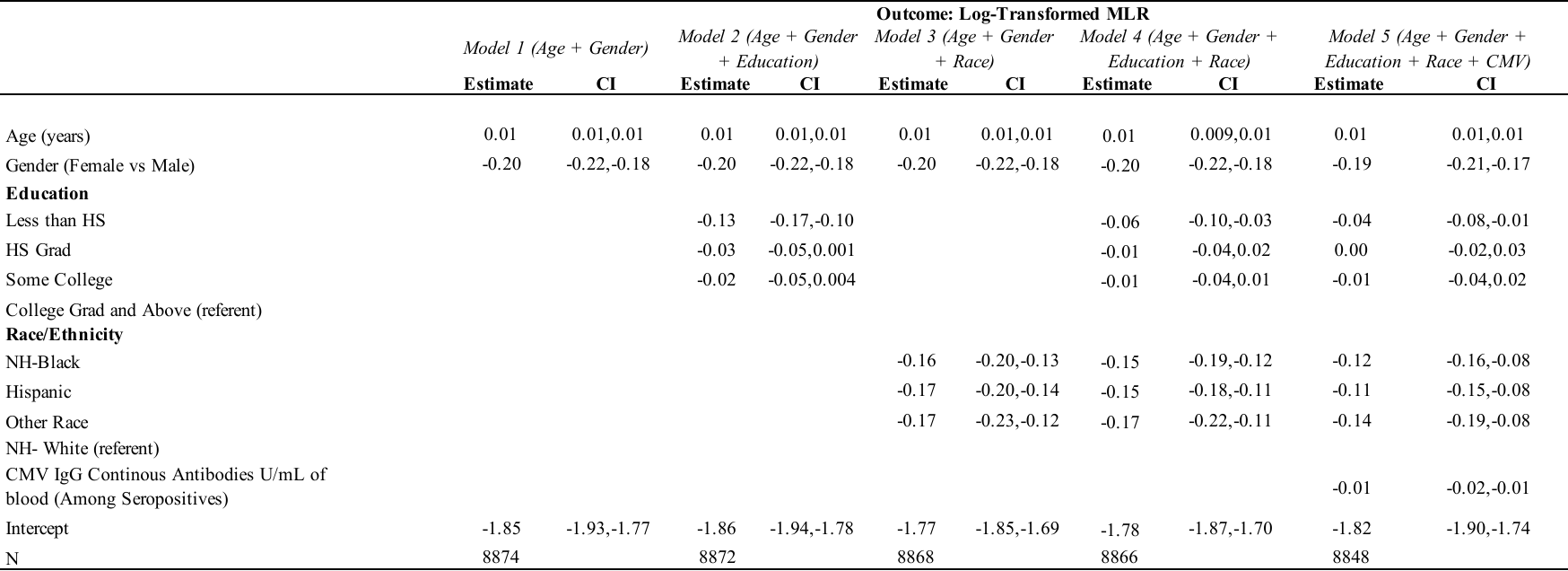

Section III: Supplemental Figures

**Figure S1. Median (95% CI) Immune Ratio Values by Sociodemographic Characteristics**
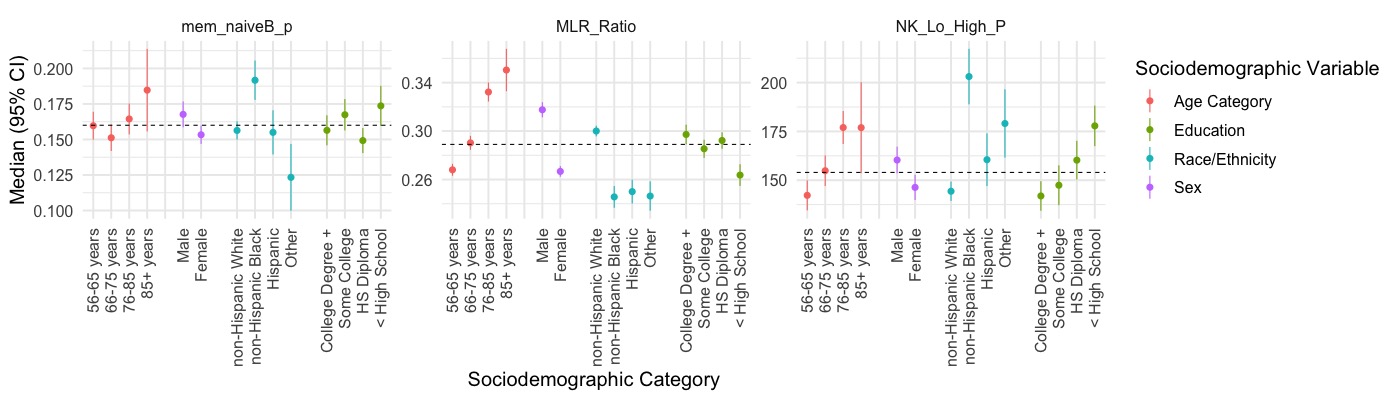

**Figure S2.A.**
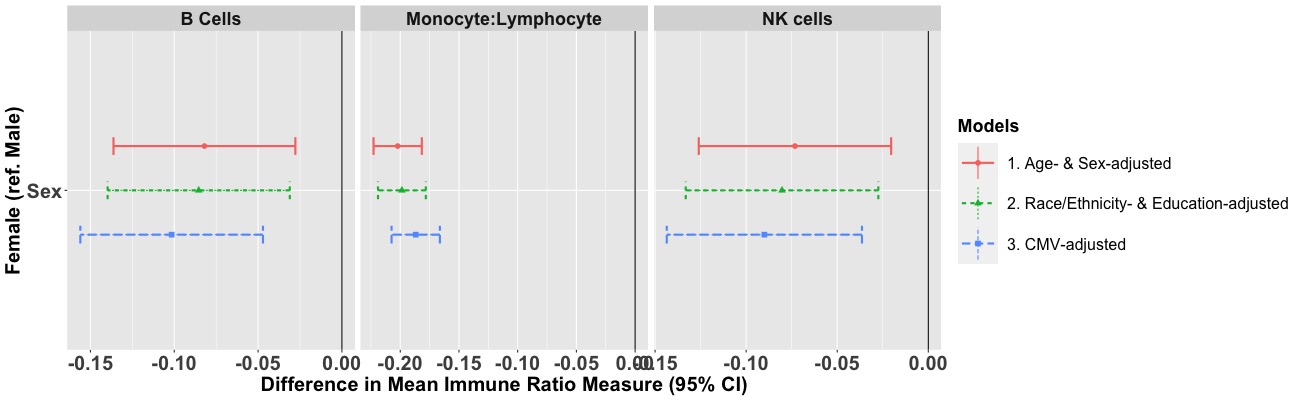

**Figure S2.B**
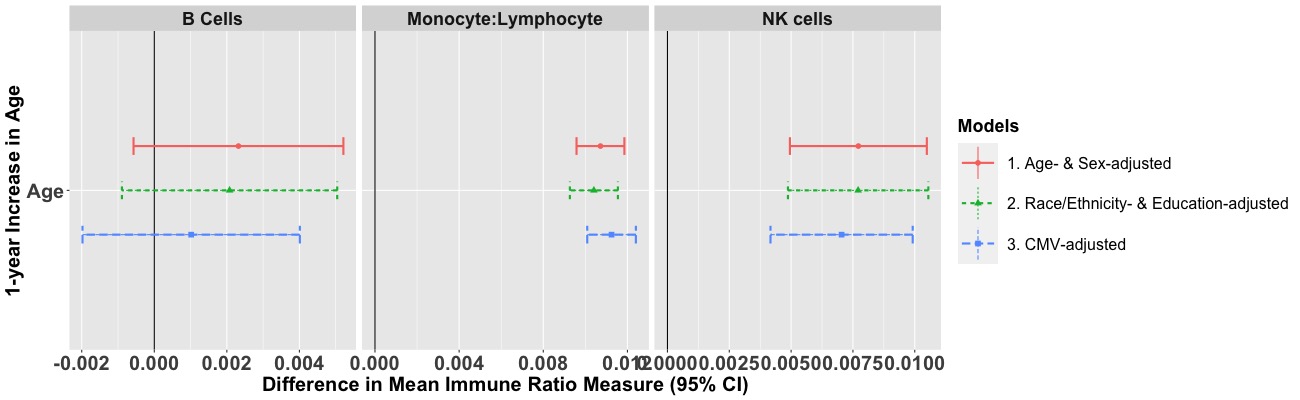

**Figure S2.C**
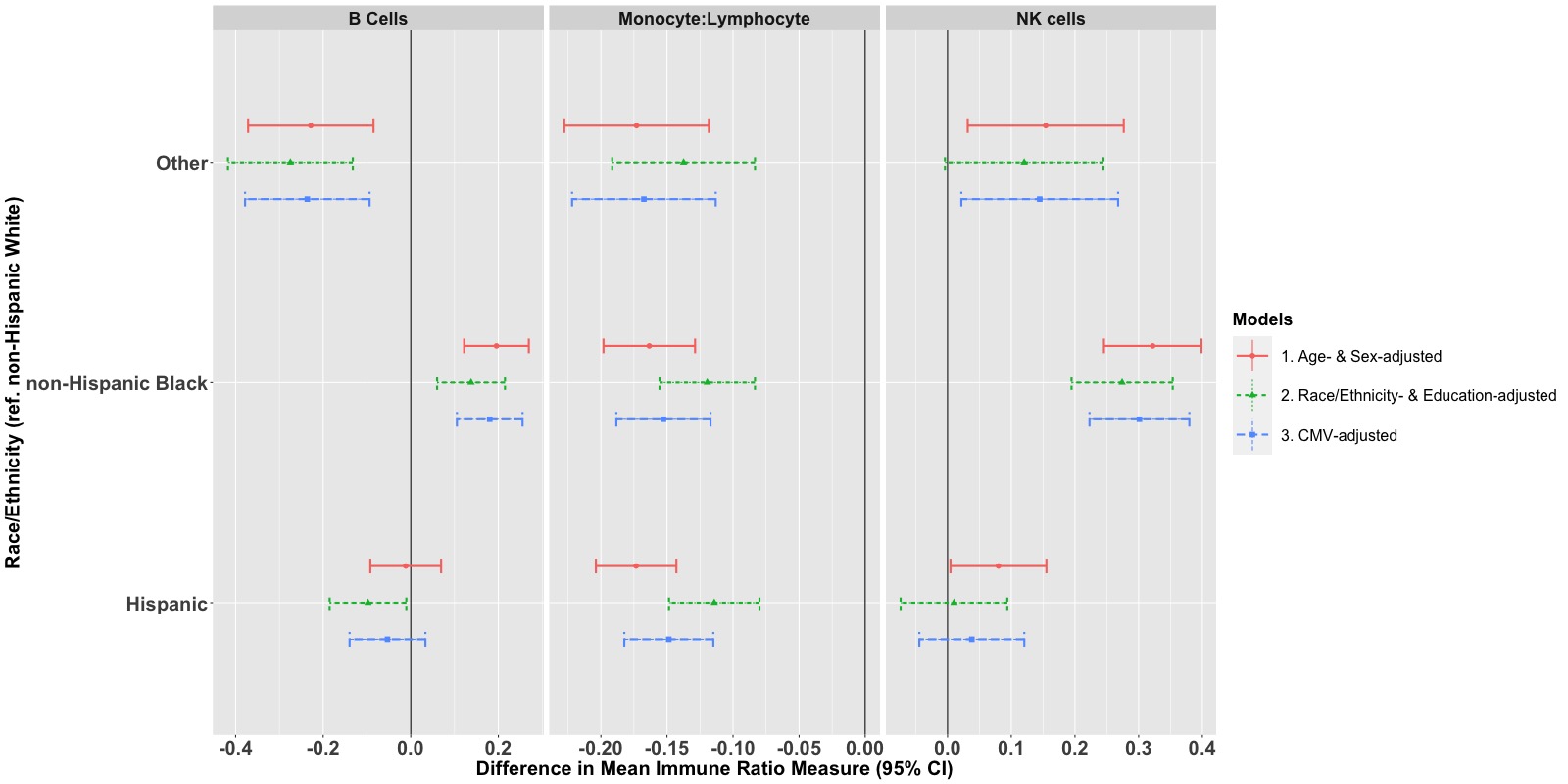
 **Figure S2.D**
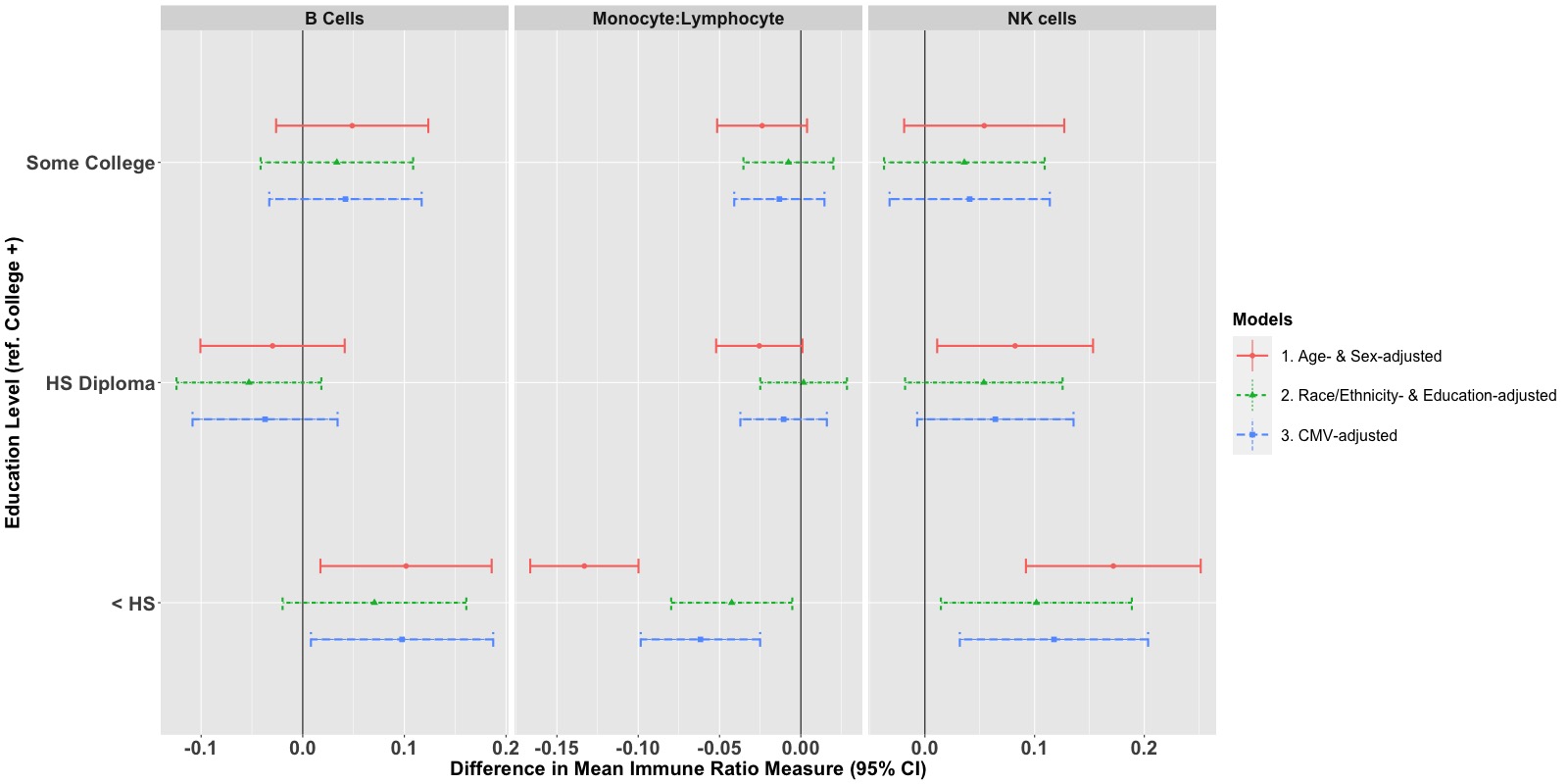

**Figure S3.**
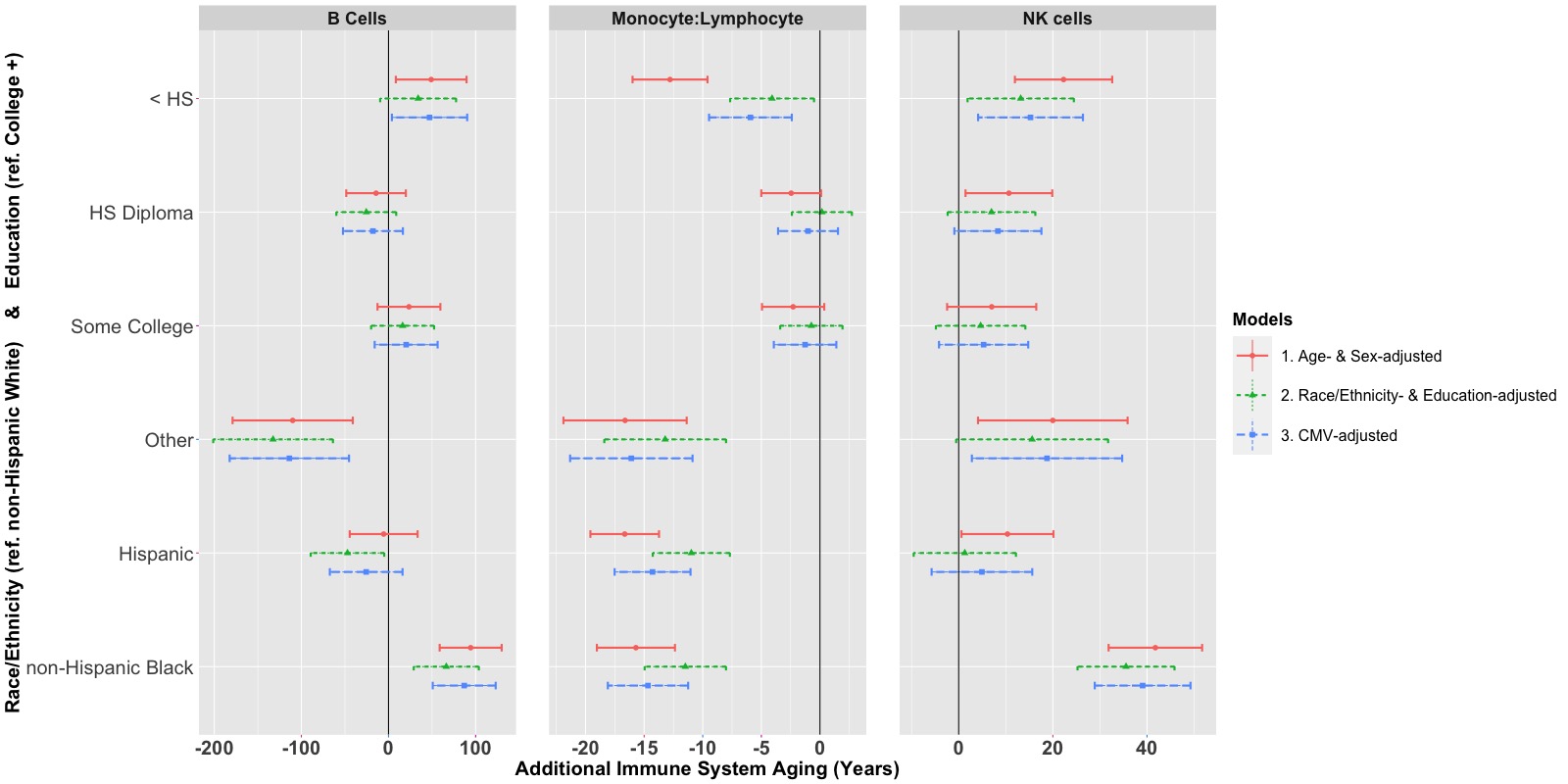
